# A cross-sectional study of quantitative CT measurements associated with the diffusion capacity of the lung in recovered COVID-19 patients with normalised chest CT

**DOI:** 10.1101/2022.09.13.22279543

**Authors:** Han Wen, Shreya Kanth, Julio Huapaya Carrera, Junfeng Sun, Michael Do, Marcus Y. Chen, Ashkan A. Malayeri, Anthony F. Suffredini

## Abstract

Impairment of the diffusion capacity of the lung for carbon monoxide (DLco) is commonly reported in convalescent and recovered COVID-19 patients, although the cause is not fully understood especially in patients with no radiological sequelae. In a group of 47 patients at 7 - 51 weeks post infection with either none or minimal scarring or atelectasis on chest CT scans (total < 0.1% of lung volume), dispersions in DLco-adj % and total lung capacity (TLC) % of predicted were observed, with median(quartiles) of 87(78, 99)% and 84(78, 92)%, respectively. Thirteen(27.1%) patients had DLco-adj% < 80%. Although the DLco-adj% did not significantly correlate with the severity of the illness in the acute phase, time since the onset of symptoms, the volume of residual lesions on CT, age or sex, DLco-adj/alveolar volume (Kco-adj) % predicted was correlated with the measurements of small blood vessel volume fraction (diameter <= 5mm) and parenchyma density on CT. Multivariate analysis revealed that these two CT metrics significantly contributed to the variance in DLco-adj% independent of TLC%. Comparing to between-subject variability of DLco-adj in healthy individuals, patients in this cohort with DLco-adj% < 80% were likely abnormal with a degree of disease not visually detectable on CT. However, it is not clear whether the associated variance of parenchyma density and small vessel volume fraction were a consequence of the COVID-19 disease or a pre-existing background variance.

---

Persistent reduction of the diffusion capacity of the lung is a frequently reported longer term consequence of COVID-19 infection with a cited prevalence of 40-65% in recovered patients(1–10). Although the reduction in DLco is often correlated with persisting radiological abnormalities(3–5, 7, 8), it has not been investigated in recovered patients with none or minimal radiologic anomalies. Here we report a cross-sectional study of 47 patients at 7 to 52 weeks post infection with normalised chest CT, defined as either none or minimal scarring or atelectasis lesions present (total volume of lesions < 0.1% of lung volume). In this group of patients, the median and quartiles of DLco adjusted for hemoglobin (DLco-adj % predicted) were 87.1(79.1, 98.4)%, of the TLC % predicted were 84.0(77.3, 91.8)%, and of the Kco-adj% predicted were 103.3(91.7, 111.6)%. Although the DLCO-adj% was not statistically correlated with clinical and demographic variables including the severity of the illness in the acute phase, time since the onset of symptoms, age, sex, or volume of residual lesions on CT (all p-values > 0.21), it was found that the Kco-adj % predicted was correlated with the quantitative CT measurements of parenchyma density (ρ = 0.343, p = 0.018) and small blood vessel volume fraction defined as (volume of vessels of diameter <= 5mm)/lung volume (ρ = 0.358, p = 0.013). When adjusted for the clinical variables mentioned above, parenchyma density and small vessel volume fraction were significant contributors to the variance of DLco-adj% (19%, p = 0.003) besides TLC% (22%, p < 0.001). These three factors accounted for 41% of the DLco-adj variance in this cohort. It is not clear whether the variance in parenchyma density and vessel volume fraction were a consequence of COVID-19 or a pre-existing background variability.

This is a retrospective study of 47 COVID-19 patients (25 male, 22 female) enrolled at the NIH Clinical Center from December 2020 to April 2022. The inclusion criteria were adult patients who received concurrent pulmonary function tests and a chest CT scan in the convalescent or recovery phase of the disease, and the chest CT was either free of any abnormalities or had residual scars and/or atelectasis totaling less than 0.1% of the lung volume. Of this cohort, 36 were Caucasian (6 Hispanic or Latino), and the median(quartiles) of age were 45.2(37.4, 56.0). 35 had mild disease in the acute phase (grade 1 to 2 on the NIAID ordinal scale), 9 were moderate (NIAID-OS of 4 to 5) and 3 were severe (NIAID-OS of 6). The mean(quartiles) of the time from the onset of first symptoms to the study date was 85(77, 267) days.

The CT exam included a volumetric high-resolution chest CT scan (HRCT). Three quantitative measurements (qCT) were obtained from the HRCT images. The first was the average parenchyma density on CT, defined as the mode of the histogram of CT pixel values (Hounsfield units) in the whole lung. The second was the small blood vessel volume fraction (diameter <= 5mm), which was obtained with a custom automated software (Fig. 1). Both measurements were adjusted to the condition of maximal inhalation based on the ratio of TLC/(total air volume in the lungs measured on CT). The third CT measurement was the total volume of residual lesions if present, based on manual segmentation of the images. Other demographic and clinical variables included in the study were age, sex, the severity of the disease on the NIAID ordinal scale, and time since the onset of first symptoms.

**Figure 1.**
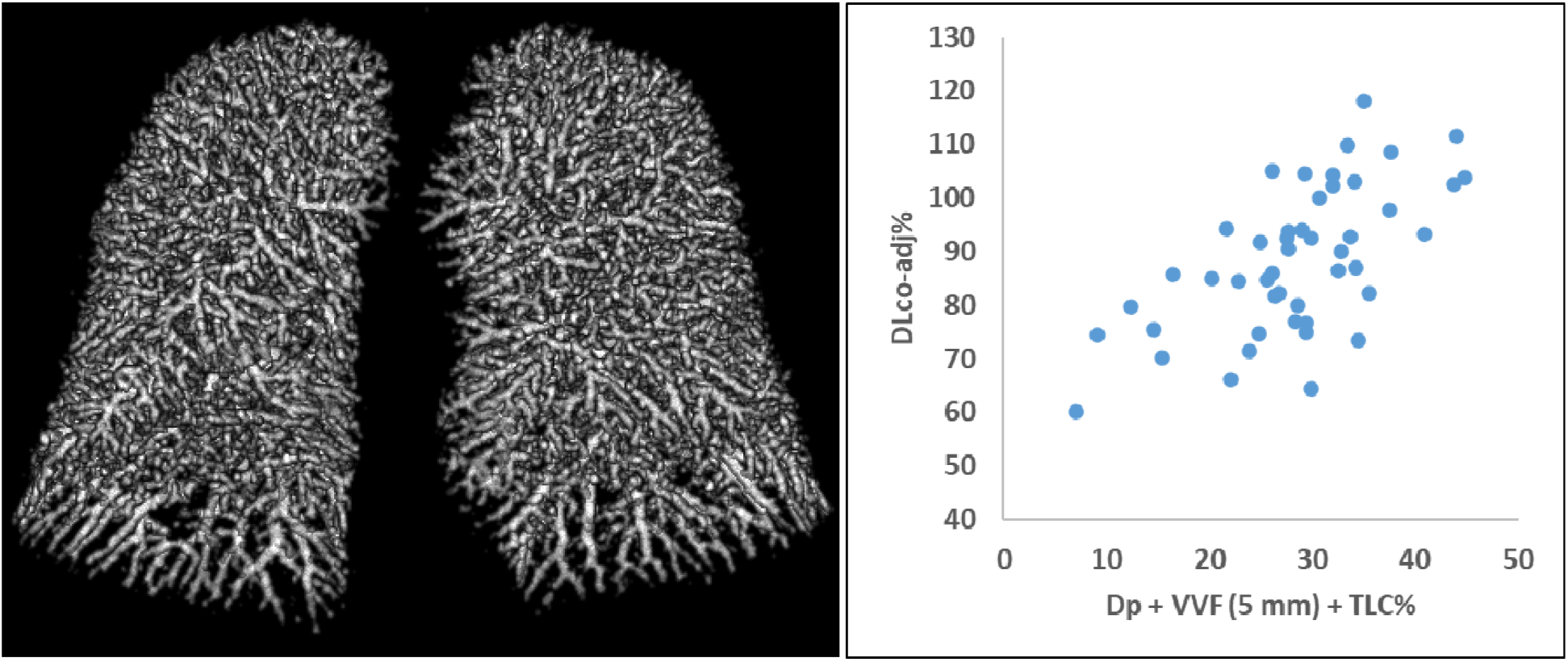
Left: 3D representation of blood vessels of diameters <= 5 mm in the lungs of a recovered COVID-19 patient. Right: A plot of DLco-adj % predicted versus a composite of the parenchyma density and small vessel volume fraction by CT, and the TLC%. These three variables explained 41% of the variance in DLco-adj among the patient cohort after adjustment for a host of demographic and clinical variables.

In the statistical analysis the outcome variables were DLco-adj and Kco-adj % predicted. The independent variables were TLC% predicted, the three quantitative CT measurements and the clinical variables. Univariate Peason’s correlation analysis showed that DLco-adj% was significantly correlated with TLC% (ρ = 0.466, p < 0.001), but not with the clinical variables (all p-values > 0.21) or the qCT measurements (all p-values > 0.20); Kco-adj% was correlated with the qCT measurements of parenchyma density (ρ = 0.343, p = 0.018) and small vessel volume fraction (ρ = 0.358, p = 0.013), but not with the clinical variables (all p-values > 0.28) or the residual lesion volume (p = 0.22). Since the two qCT measurements of parenchyma density and small vessel volume fraction were mutually correlated (ρ = 0.569, p < 0.001), we used principal component analysis to extract the two independent components, namely PC_1_ representing a common covariate of the two, and PC_2_ representing the difference between the two. The common covariate PC_1_ was correlated with Kco-adj% (ρ = 0.396, p < 0.006), but not the difference PC_2_ (ρ = -0.032, p = 0.83).

We used a multiple linear regression analysis to determine 1. whether the parenchyma density and small vessel volume fraction on CT contributed to the variance in DLco-adj when adjusted for the clinical and demographic variables, and 2. whether their contribution was independent of that of the TLC%. The result was that besides TLC%, only the covariate component of the two qCT metrics (PC_1_) was a statistically significant contributor (p < 0.003). The difference component PC_2_ did not reach statistical significance (p = 0.14), nor did any of the demographic and clinical variables (p-values > 0.3). The parenchyma density and small vessel volume fraction together accounted for 19% of the variance in DLco-adj% predicted, and TLC% accounted for another 22%. The combination of these three factors accounted for 41% of the DLco-adj variance (Fig. 1).

In this cohort, the alveolar volume was highly correlated with the TLC volume (ρ = 0.966). Previous studies have debated whether a lingering low DLco after recovery from COVID-19 is mainly due to reduced alveolar volume and not interstitial or vascular abnormalities(11), or involves the presence of abnormal perfusion or gas exchange(6, 12). The data from the current study suggests that in recovered patients with normalised chest CT, there is a variance of the lung parenchyma and vascular structure that contributes to the dispersion in DLco, and the contribution is independent from the alveolar volume. However, without pre-COVID measurements it is not clear whether this variance of the tissue structure is a consequence of COVID-19 or pre-existing background variability(13). This is the main limitation of the current study.

## Data Availability

All data produced in the present work are contained in the manuscript.

